# Sarcomere gene variants did not improve cardiac function in pediatric patients with dilated cardiomyopathy from Japanese cohorts

**DOI:** 10.1101/2024.01.24.24301754

**Authors:** Keiichi Hirono, Yukiko Hata, Shojiro Ichimata, Naoki Nishida, Teruhiko Imamura, Yoshihiro Asano, Yuki Kuramoto, Kaori Tsuboi, Shinya Takarada, Mako Okabe, Hideyuki Nakaoka, Keijiro Ibuki, Sayaka Ozawa, Jun Muneuchi, Kazushi Yasuda, Kotaro Urayama, Hideharu Oka, Tomoyuki Miyamoto, Kenji Baba, Akio Kato, Hirofumi Saiki, Naoki Kuwahara, Masako Harada, Shiro Baba, Mari Morikawa, Hidenori Iwasaki, Yuichiro Hirata, Yuki Ito, Heima Sakaguchi, Susumu Urata, Koichi Toda, Emi Kittaka, Seigo Okagda, Yohei Hasebe, Shinsuke Hoshino, Takanari Fujii, Norie Mitsushita, Masaki Nii, Kayo Ogino, Mitsuhiro Fujino, Yoko Yoshida, Yutaka Fukuda, Satoru Iwashima, Kiyohiro Takigiku, Yasushi Sakata, Ryo Inuzuka, Jun Maeda, Yasunobu Hayabuchi, Tao Fujioka, Hidemasa Namiki, Shuhei Fujita, Koichi Nishida, Ayako Kuraoka, Nobuhiko Kan, Sachiko Kido, Ken Watanabe, Fukiko Ichida

**Affiliations:** Department of Pediatrics, Faculty of Medicine, University of Toyama, Toyama, Japan; Legal Medicine, Faculty of Medicine, University of Toyama, Toyama, Japan; 2^nd^ department of Internal Medicine, Faculty of Medicine, University of Toyama, Toyama, Japan; National Cerebral and Cardiovascular Center, Osaka, Japan; Department of Cardiovascular Medicine, Osaka University Graduate School of Medicine, Osaka Japan; Department of Pediatrics, Kyushu Hospital, Japan Community Healthcare Organization, Fukuoka, Japan; Department of Pediatric Cardiology, Aichi Children’s Health and Medical Center, Aichi, Japan; Department of Pediatrics, Tsuchiya General Hospital, Hiroshima, Japan; Department of Pediatrics, Asahikawa Medical University Hospital, Hokkaido, Japan; Department of Pediatrics, Yokosuka General Hospital, Kanagawa, Japan; Department of Pediatrics Okayama University Graduate School of Medicine, Okayama, Japan; Department of Pediatric Cardiology, Okinawa Prefectural Nanbu Medical Center and Children’s Medical Center, Okinawa, Japan; Department of Pediatrics and Pediatric Cardiology, Iwate Medical University School of Medicine, Iwate, Japan; Department of Pediatric Cardiology, Gifu prefectural General Medical Center, Gifu, Japan; Division of Pediatrics, Developmental and Urological-Reproductive Medicine, Faculty of Medicine, University of Miyazaki, Miyazaki, Japan; Department of Pediatrics, Kyoto University Hospital, Kyoto, Japan; Department of Pediatrics, Kanazawa Medical University, Ishikawa, Japan; Department of Pediatrics, Kanazawa University Hospital, Ishikawa, Japan; Department of Pediatrics, Kyushu University Hospital, Fukuoka, Japan; Department of Pediatric Cardiology, National Cerebral and Cardiovascular Center, Osaka, Japan; Division of Cardiology, National Center for Child Health and Development, Tokyo, Japan; Department of Pediatric Cardiology, Saitama Medical University International Medical Center, Saitama, Japan; Department of Cardiology, Saitama Children’s Medical Center, Saitama, Japan; Department of Pediatrics, Yamaguchi University Graduate School of Medicine, Yamaguchi, Japan; Department of Pediatrics, Faculty of Medicine, University of Yamanashi, Yamanashi, Japan; Department of Pediatrics, Shiga University of Medical Science, Shiga, Japan; Pediatric Heart Disease and Adult Congenital Heart Disease Center, Showa University Hospital, Tokyo, Japan; Department of Cardiology, Shizuoka Children’s Hospital, Shizuoka, Japan; Kurashiki Central Hospital, Department of Pediatrics, Okayama, Japan; Department of Pediatric Cardiology, Osaka city general hospital, Osaka, Japan; Decision of pediatric electrophysiology, Osaka city general hospital, Osaka, Japan; Department of Pediatrics, Takeda General Hospital, Fukushinma, Japan; Department of Pediatrics, Chutoen General Medical Center, Shizuoka, Japan; Department of Pediatric Cardiology, Nagano Children’s hospital, Nagano, Japan; Division of Pediatrics and Perinatology, Faculty of Medicine, Tottori University, Tottori, Japan; Department of Pediatrics, Tokyo University Hospital, Tokyo, Japan; Division of Cardiology, Tokyo Metropolitan Children’s Medical Center, Tokyo, Japan; Department of Pediatrics, Tokushima University Hospital, Tokushima, Japan; Department of Pediatrics, Japanese Red Cross Medical Center, Tokyo, Japan; Department of Pediatrics and Child Health, Nihon University School of Medicine & Itabashi Hospital, Tokyo, Japan; Department of Pediatrics, Toyama Prefectural Hospital, Toyama, Japan; Department of Pediatrics, Fukui Cardiovascular Center, Fukui, Japan; Department of Cardiology, Fukuoka Children’s Hospital, Fukuoka, Japan; Department of Fetal and Neonatal Cardiology, Fukuoka Children’s Hospital, Fukuoka, Japan; Department of Cardiology, Hyogo Prefectural Kobe Children’s Hospital, Kobe, Japan; Department of Pediatrics, Kitano Hospital Tazuke Kofukai Medical Research Institute, Osaka Japan; Department of Pediatrics, International University of Health and Welfare, Tokyo, Japan

**Keywords:** dilated cardiomyopathy, heart failure, genetics, sarcomere gene, left ventricular reverse remodeling

## Abstract

**Background:** Dilated cardiomyopathy (DCM) is a progressive myocardial disorder characterized by impaired cardiac contraction and ventricular dilation. Some patients with DCM could manifest improvement in these abnormalities called left ventricular reverse remodeling (LVRR). However, the detailed association between genotypes and clinical outcomes, including LVRR, particularly among pediatric patients, remains uncertain.

*Methods:* We prospectively enrolled pediatric patients with DCM from Japanese multi-institutional centers between 2014 and 2023. We identified DCM-related genes and explored the association between gene variants and clinical outcomes, including LVRR, which was defined as any increase in left ventricular ejection fraction during the observation period.

*Results:* A total of 123 pediatric patients (62 males; mean age of 8 months [range, 1–51 months]) were retrospectively enrolled. There were 50 pathogenic variants in 45 patients (35.0%). The most identified gene was *MYH7* (14.0%), followed by *RYR2* (12.0%), and *TPM1* (8.0%). A novel variant in the *CASZ1* gene (NM_001079843.2 c.3356G>A, p. Trp1119Ter) was identified. LVRR was achieved in 47.5% of patients. In patients with sarcomere gene variants, the left ventricular ejection fraction remained unchanged (31.4% to 39.8%, *P* = 0.1913), whereas it significantly increased in patients with non-sarcomere gene variants (33.4% to 47.8%, *P* = 0.0466) and in patients without gene variants (33.6% to 54.1%, *P* = 0.003).

*Conclusions:* Pediatric patients with DCM exhibited a marked genetic heterogeneity with a different landscape from adults with DCM. LVRR was not uniform across functional gene groups, opening the door to tailor-made gene-guided prediction in pediatric patients with DCM.

*Clinical perspective:* What is new?

- Younger patients had predominance for DCM and risk factors for survival.
- LVRR occurrence was lower in the sarcomere gene group, and cardiac function failed to improve. What are the clinical implications?

- LVRR was not uniform across functional gene groups, which opens the door to the adoption of an individualized prediction approach in DCM according to the genetic features.

## INTRODUCTION

Dilated cardiomyopathy (DCM) is a myocardial disorder with the potential for heart failure (HF) and fatal outcomes. It is characterized by impaired cardiac contraction and dilation of the left ventricle (LV) or both ventricles without obvious etiologies. In both adult and pediatric populations, DCM is a major cause of myocardial diseases. In adults, the prevalence of DCM is estimated to be 1 in 2,500.^1,2^ Conversely, in the pediatric population, the annual incidence rate is considerably lower, with estimates of one in 170,000 in the United States (U.S.), one in 140,000 in Australia, and one in 200,000 in Japan.^3,4^ While the annual incidence of pediatric DCM is lower than that in adults, the clinical outcomes of pediatric DCM patients can be particularly severe. Moreover, life-threatening cardiac outcomes was recorded to be 42% for heart transplants and account for 5% of deaths in the U.S.^5,6^

The occurrence of multiple DCM in patients within a single pedigree indicates genetic contributions.^7,8^ Recent reports indicate that approximately 30%–40% of DCM cases are caused by pathogenic gene variants, with over 50 genes suggested to be associated with the condition.^1,9,10^ The pathophysiology of DCM involving genetic factors is linked to genes encoding sarcomere proteins, cellular cytoskeleton components, nuclear membrane proteins, and ion channels. Nevertheless, most genetic studies have been limited to adults. Consequently, understanding the genetic causes of primary DCM that occurs during childhood is reliant on limited research. Despite recommendations for genetic testing in pediatric cardiomyopathy, variability in clinical practice exists because of the lack of larger-scale studies in children.

Heart remodeling in response to myocardial stress has traditionally been considered a characteristic feature of DCM. Recently, several cohort studies have demonstrated that in a subset of patients with DCM, a process generally referred to as left ventricular reverse remodeling (LVRR) can lead to the reversal of this phenomenon. These findings suggest that DCM may not follow an irreversible progression of myocardial damage but rather represent a reversible and dynamic disease if correctly treated.^11,12^ Several studies have also reported successful LVRR in pediatric DCM patients.^13–15^ However, specific genotypes associated with LVRR in children have not been elucidated.^11^

As genetic data on pediatric cardiomyopathy are limited, this study aimed to investigate the genetic structure of pediatric-onset DCM through a nationwide survey in Japan. Additionally, this study examined the association between disease-causing genes and LVRR.

## METHODS

### Study population

From October 2014 to March 2023, a total of 123 Japanese probands with DCM were referred to our institution from 45 Japanese hospitals for genetic testing. The patients were aged <18 years and were recently diagnosed with DCM. The exclusion criteria included children with secondary cardiomyopathies (e.g., neuromuscular, pulmonary, endocrine, rheumatic, and immunologic diseases; inborn errors of metabolism with multiple organ involvement; and significant structural heart disease), cardiotoxic exposures, systemic hypertension, and missing follow-up records.

Clinical data were retrospectively retrieved from the patients’ medical records. The following clinical characteristics were collected during the initial presentation: (1) presence and nature of cardiac symptoms, (2) reason for referral, (3) presence of concomitant cardiac or extracardiac involvements, and (4) family history. Cardiac death, LV assist device implantation, heart transplantation, and appropriate implantable cardioverter-defibrillator (ICD) shock were classified as major adverse cardiovascular events (MACE).

According to institutional guidelines, informed consent was obtained from all patients or their parents. The study protocol conformed to the ethical guidelines of the 1975 Declaration of Helsinki, as reflected by the a priori approval of the Research Ethics Committee of the University of Toyama, Japan.

### Electrocardiogram collection

A 12-lead standard electrocardiogram (ECG) at a speed of 25 mm/s and a voltage of 10 mm/mV (filter range 0.15–100 Hz; AC filter: 60 Hz) was obtained. Two well-trained investigators (K.H. and N.M.) blinded to clinical data independently evaluated all ECGs. More than 95% of the first judgments were consistent between the two investigators. In cases where there was a disagreement, a third experienced investigator (electrophysiologist) would make the final judgment (two-third agreement constitutes the decision).

### Echocardiographic data

All echocardiographic data were analyzed by two independent reviewers (K.H. and S.O.) who were blinded to each other’s measurements. Echocardiography (2D, M-mode, and color Doppler) was used to evaluate cardiac structure, LV size and function (left ventricular ejection fraction [LVEF]), and valve regurgitation. The thickness of the compacted layer in the LV and LV diastolic diameter (LVDD) were expressed as Z-scores, which were indexed to the body surface area.^16^

DCM was defined as LV dilation with a decreased LV systolic function.^17,18^ LV dilation was defined as an LV end-diastolic dimension more than 2 standard deviations above the mean normal value for body surface area (LVDD *Z* score >2), and LV systolic dysfunction was defined as an LVEF more than 2 standard deviations below the mean value for healthy children, which is the adjusted normal value for age (LVEF Z score of <2, <50 %).^17,18^

Given the retrospective nature of this study, we first identified patients diagnosed with DCM and then reviewed their echocardiograms to ensure that they met all inclusion criteria. HF was diagnosed based on clinical symptoms of feeding difficulty and tachypnea, decreased LVEF on echocardiography, and cardiomegaly on chest X-ray. Cardiomegaly was defined as a cardiothoracic ratio of ≥0.55 (≥0.60 for patients <1 year old) on chest X-ray.

LVRR was defined as either LV normalization (LVEF improvement to ≧50% with a ≧5% LVEF increment on echocardiogram at 1 year after) or an absolute increase in LVEF by ≧10% on echocardiogram at 1 year after from initial echocardiogram at baseline, as described.^19,20^

### Genetic testing using next-generation sequencing (NGS)

Genomic DNA was extracted from whole blood samples obtained from the patients using the QIAamp DNA Mini Kit (Qiagen, Redwood City, CA, USA). NGS of 203 cardiovascular disease-related genes associated with cardiomyopathies and channelopathies (Table S1) was conducted using the Ion PGM System (Thermo Fisher Scientific, Waltham, MA, USA). This custom panel used two separate polymerase chain reaction (PCR) primer pools, yielding a total of 6,138 amplicons and was used to generate target amplicon libraries. Genomic DNA samples were PCR-amplified using a custom panel and an Ion AmpliSeq Library Kit Plus (Thermo Fisher Scientific). Individual samples were labeled using the Ion Xpress Barcode Adapters Kit (Thermo Fisher Scientific) and then pooled at equimolar concentrations. Emulsion PCR and ion sphere particle enrichment were performed using the Ion PGM™ Hi-Q™ View OT2 Kit (Thermo Fisher Scientific) according to the manufacturer’s instructions. Ion sphere particles were loaded onto a 318 chip and sequenced using the Ion PGM™ Hi-Q™ View Sequencing Kit (Thermo Fisher Scientific). The Torrent Suite and Ion Reporter Software version 5.0 (Thermo Fisher Scientific) were used to perform primary, secondary, and tertiary analyses, including the optimized signal processing, base calling, sequence alignment, and variant analysis.

### Variant filtration

For subsequent filtering, minor allele frequency was estimated using the Genome Aggregation Database (gnomAD) and the Japanese allele frequency in the Tohoku University Tohoku Medical Megabank Organization (Japanese Multi-omics Reference Panel [jMorp]; https://jmorp.megabank.tohoku.ac.jp). Filter impact ratings of the high (start loss, stop gain, stop loss, frameshift, and splice acceptor/donor) and moderate (missense and in-frame insertion/deletion and protein-altering) variants were then employed at a frequency of <0.0005 in gnomAD, Tohoku University Tohoku Medical Megabank Organization 8.3KJPN (ToMMo 8.3KJPN), and a CADD score ≥20.

### Selection of potential variants

To investigate the pathogenicity of the possible variants, we used 5 different *in silico* predictive algorithms: FATHMM, SIFT, Align GVGD, MutationTaster2, and PolyPhen-2 (Table S2). Subsequently, only the variants with all “pathogenic” at 4 of the 5 in silico tools were selected.

### Classification of pathogenicity

The variants were manually evaluated according to the detailed information obtained from ClinVar (https://www.ncbi.nlm.nih.gov/clinvar/) and the Human Gene Mutation Database (http://www.hgmd.cf.ac.uk/ac/index.php). They were classified according to the American College of Medical Genetics and Genomics guidelines.

### Sanger sequencing

For all candidate pathogenic variants that satisfied the selection criteria, Sanger sequencing was used to validate the NGS results. Furthermore, the nucleotide sequences of the amplified fragments were analyzed through direct sequencing in both directions using the BigDye Terminator v3.1 Cycle Sequencing Kit (Applied Biosystems, Foster City, CA). Sequence analysis was performed using an ABI 3130xl automated sequencer (Applied Biosystems).

### Statistical analysis

Continuous variables with normal distribution were expressed as mean ± standard deviation, and those with nonnormal distribution were expressed as median with interquartile range. Conversely, categorical variables were expressed as numbers and frequencies (%). To compare continuous variables, an unpaired *t*-test, nonparametric Mann–Whitney U test, or one-way analysis of variance was used, and χ^2^ statistics or Fisher’s exact test was used for the categorical variables.

Time-to-event data were presented as Kaplan–Meier estimates and compared using the log-rank test. Baseline variables that were considered clinically relevant or showed significant univariate relationships with adverse events were included in the multivariable Cox proportional hazards regression models. Variables to be included were carefully selected, considering the number of events, to ensure parsimony of the final models. Statistical analyses were conducted using JMP software (version 17; SAS institute, Cary, NC, USA). To determine the optimal cutoff values of the number of derivations obtained from chest X-ray and echocardiographic data for predicting MACE and hospitalization, receiver operating characteristic curve analysis was performed. *P* < 0.05 was considered statistically significant.

## RESULTS

### Baseline clinical characteristics

A total of 123 patients (62 males and 61 females; mean age, 8 months [range, 1–51 months]) were enrolled in this study (Table 1). The follow-up duration ranged from 4.0 to 58 months (median, 25 months). The distribution of age at onset is presented in Figure 1. Most patients were diagnosed under 1 year of age, and a minimal peak was observed during school-age.

**Figure 1.**
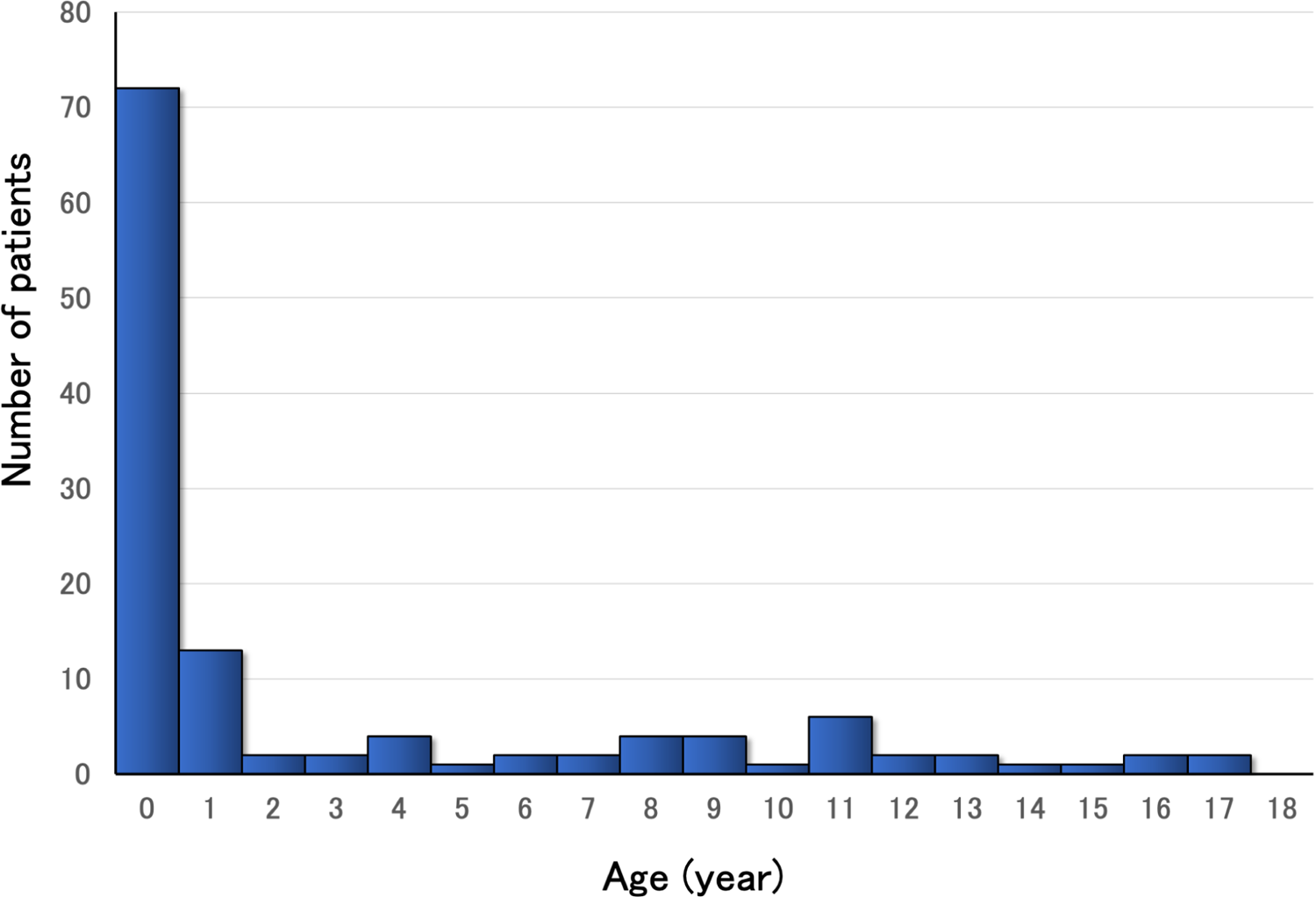
Age distribution of the patients.

**Table 1.**
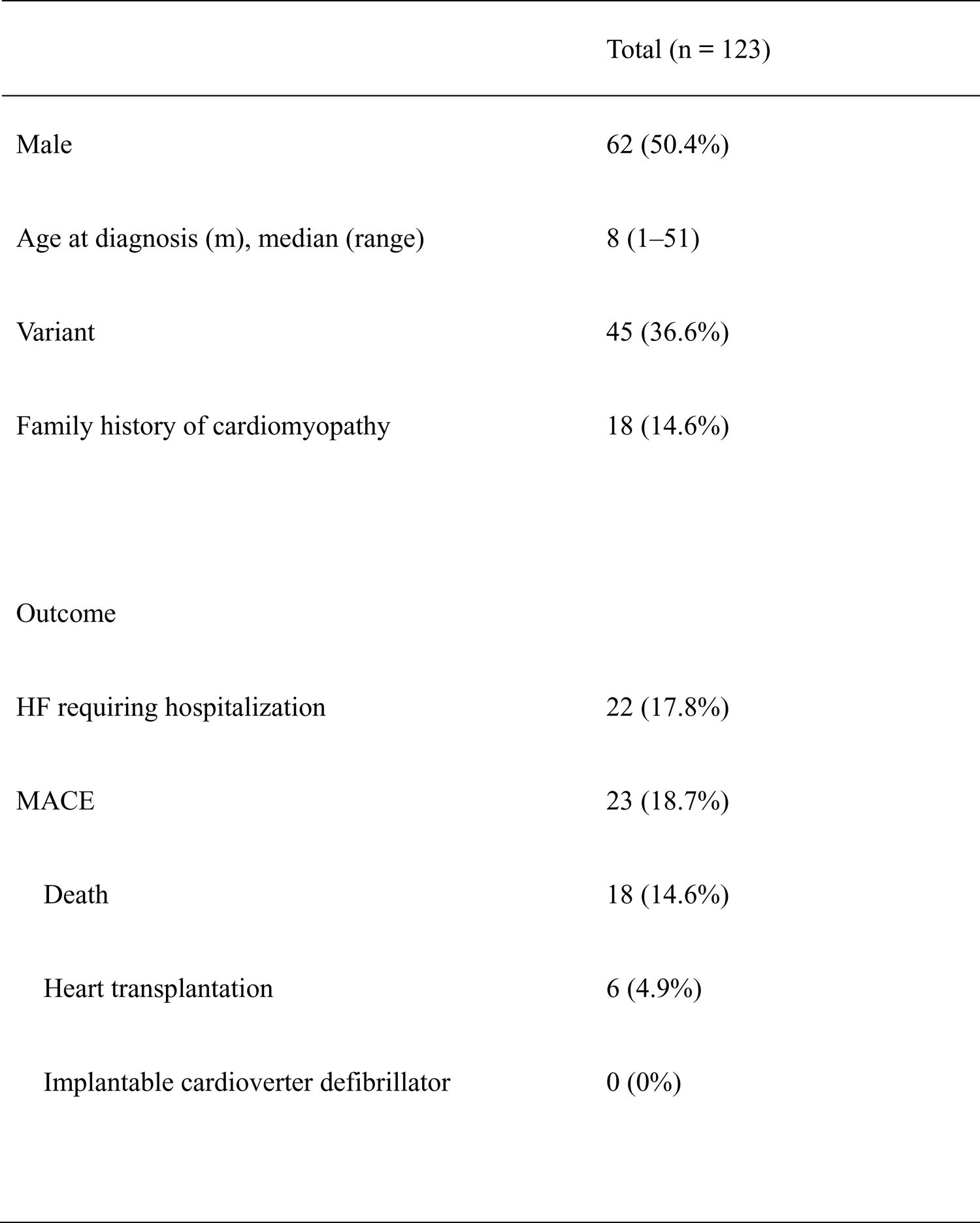
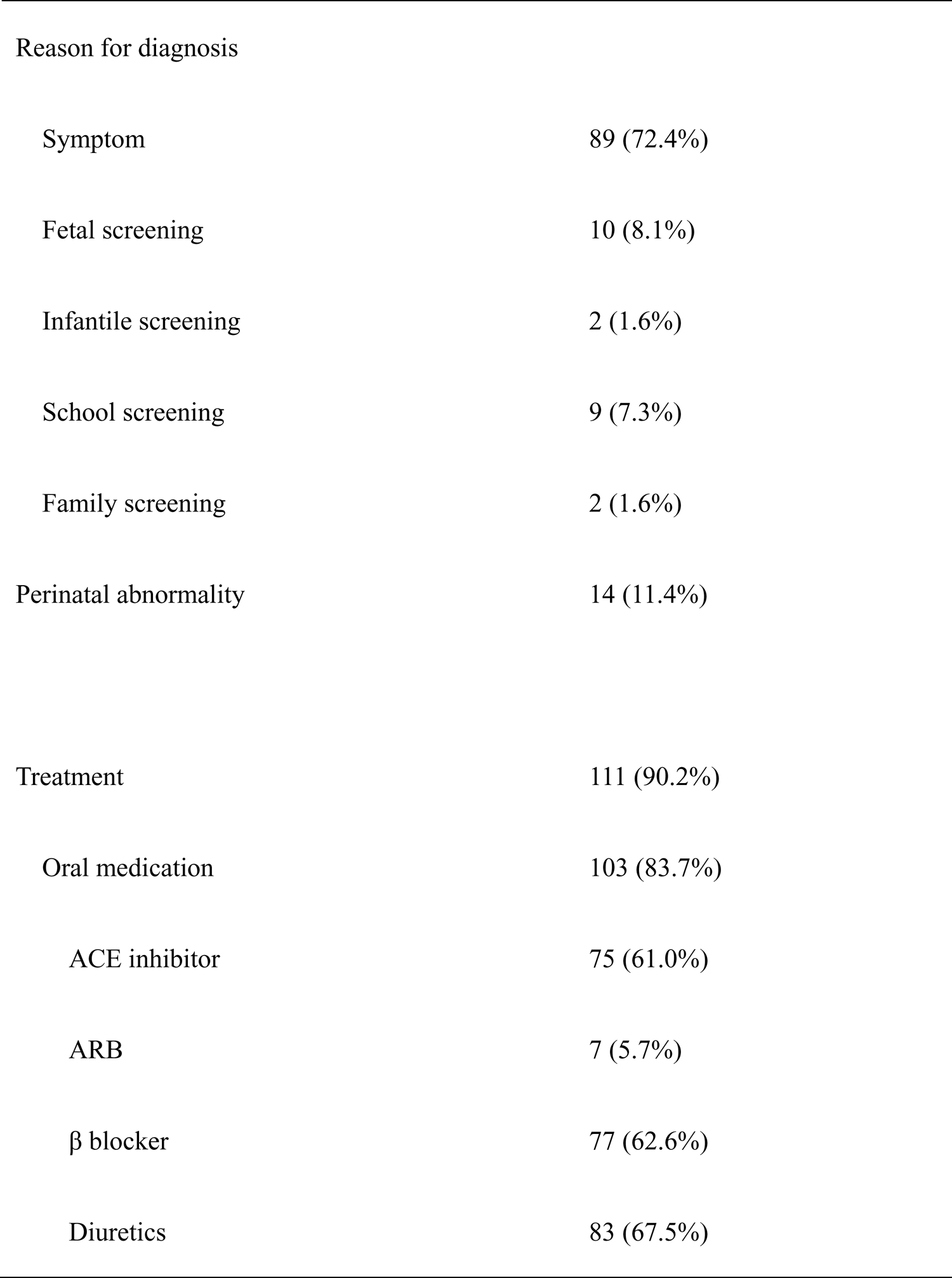
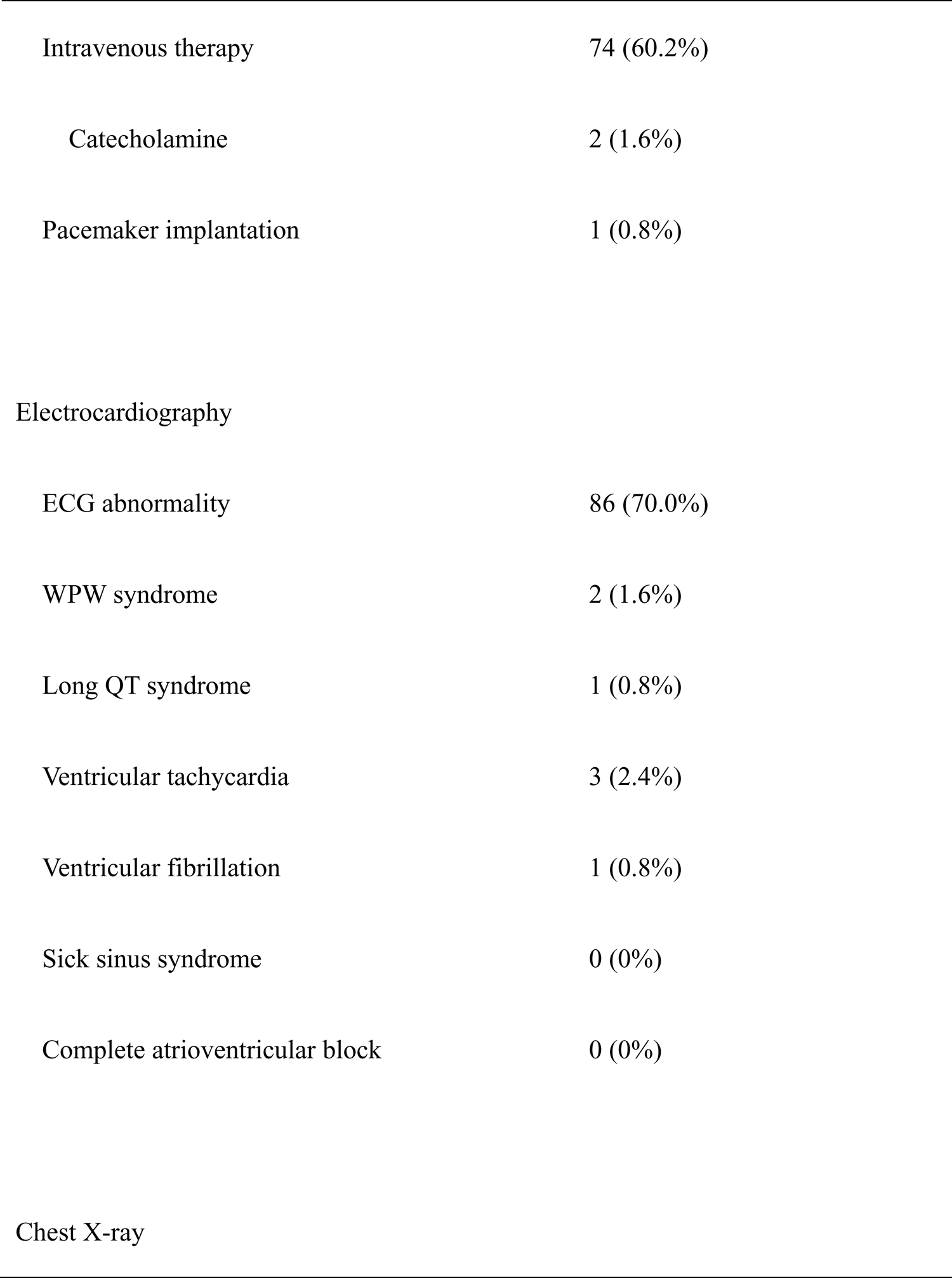
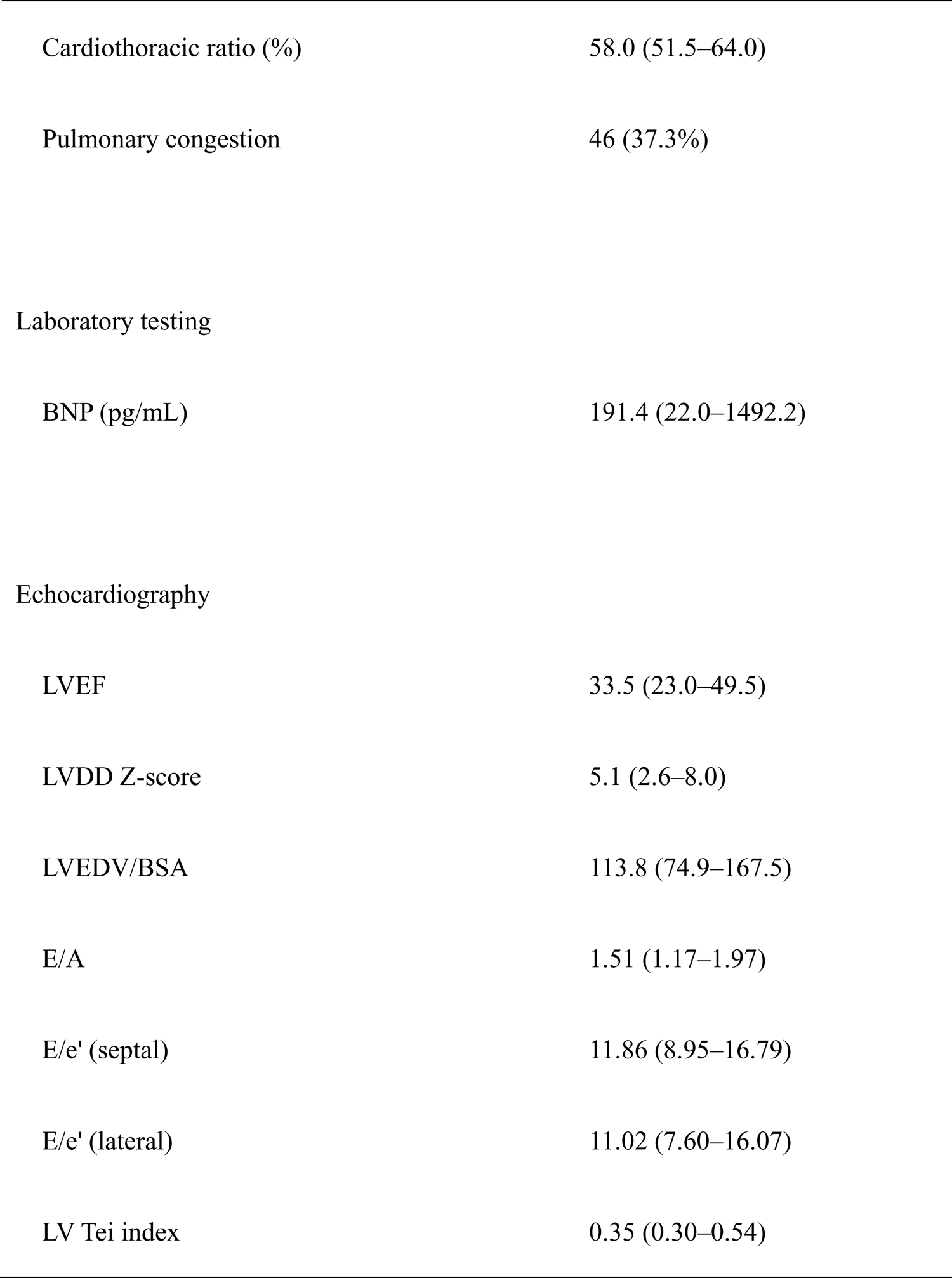
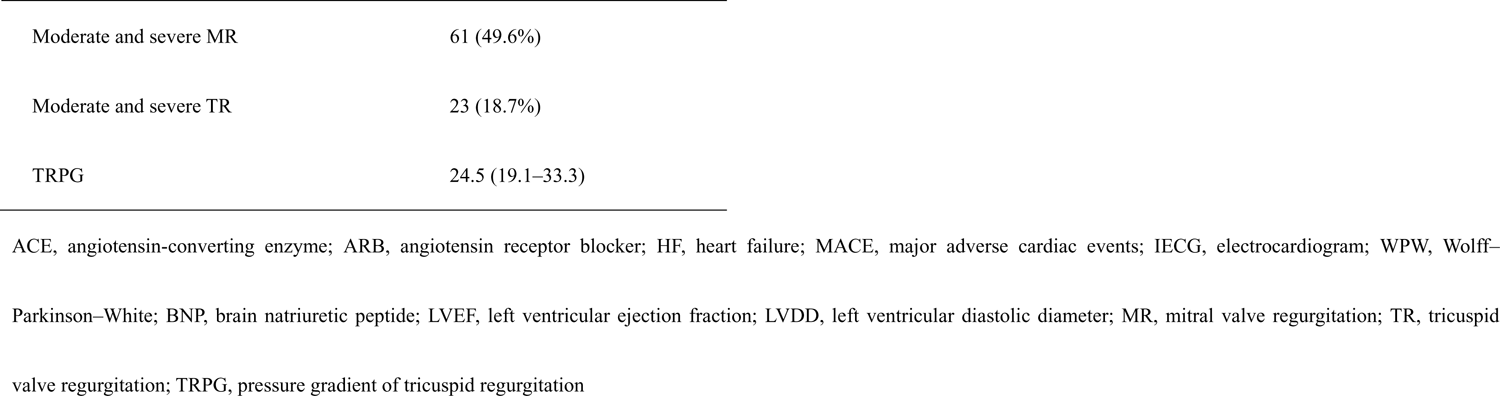
Baseline characteristics.

Detailed characteristics of the patients are presented in Table 1. Patients were diagnosed through clinical symptoms in 89 patients (72.4%) and fetal screening in 10 patients (8.1%). Perinatal abnormalities were observed in 14 patients (11.4%). Furthermore, 86 patients (70.0%) manifested ECG abnormalities. Ventricular tachycardia was observed in three patients (2.4%) and ventricular fibrillation in one patient (0.8%). Supraventricular tachycardia or bradycardia, such as complete atrioventricular block and sick sinus syndrome, was not observed.

A high cardiothoracic ratio and pulmonary congestion on chest X-ray were frequently observed. The plasma brain natriuretic peptide (BNP) level was high in patients with DCM (191.4 pg/mL [22.0–1492.2 pg/mL]). Moreover, a reduced LVEF (33.5% [23.0%–49.5%]) and high Z-scores in LVDD (5.1 [2.6–8.0]) were common.

Sequential changes in LVEF and LVDD between baseline and follow-up were observed. LVEF at follow-up was significantly increased compared with that at baseline (33.5% [23.0%– 49.5%] to 54.0% [28.2%–67.9%], *P* < 0.0001). Z-scores of LVDD at follow-up were significantly decreased compared with that at baseline (5.1 [2.6–8.0] to 1.8 [0.9–4.7], *P* < 0.0001).

### Genetic and phenotypic analyses

The distribution of the pathogenic variants is presented in Table 2. There were 50 pathogenic variants in 45 patients (35.0%) with DCM: 41 missense, 3 deletions, 5 nonsense, and 1 splicing variants. The most commonly identified gene was *MYH7* (n = 7, 14.0%), followed by *RYR2* (n = 6, 12.0%), and *TPM1* (n = 4, 8.0%). Multiple variants were identified in 5 patients (4.1%).

**Table 2.**
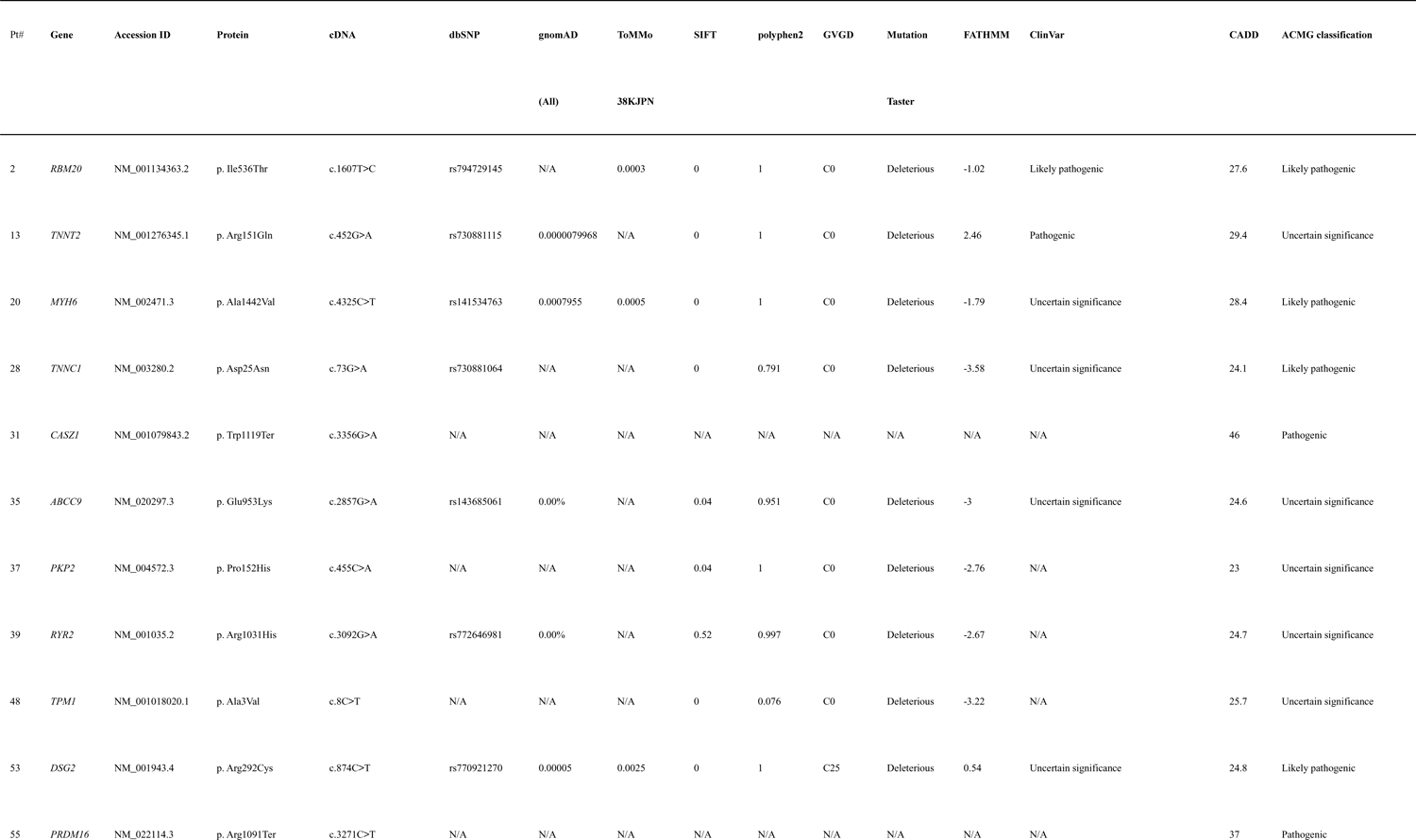
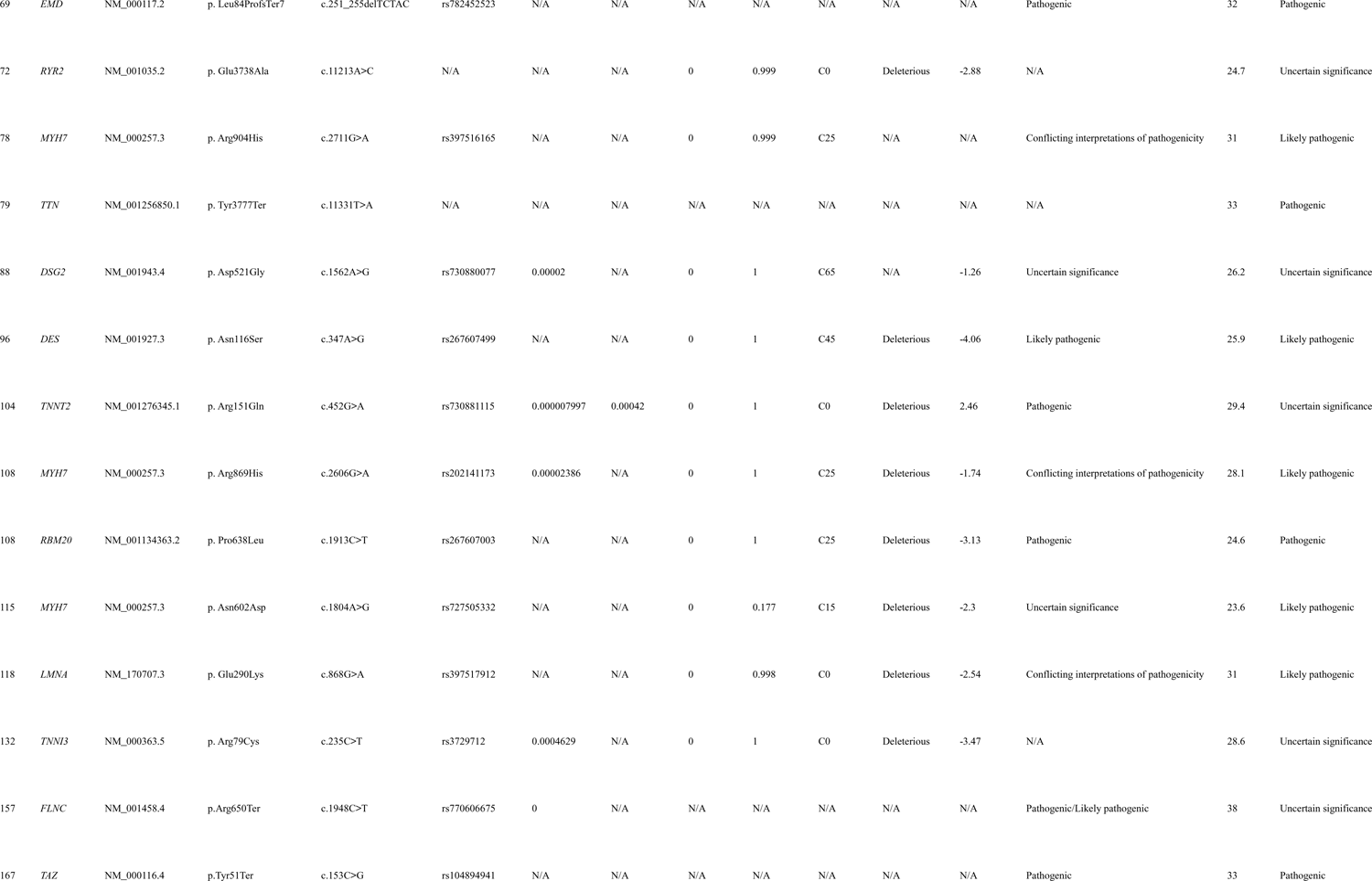
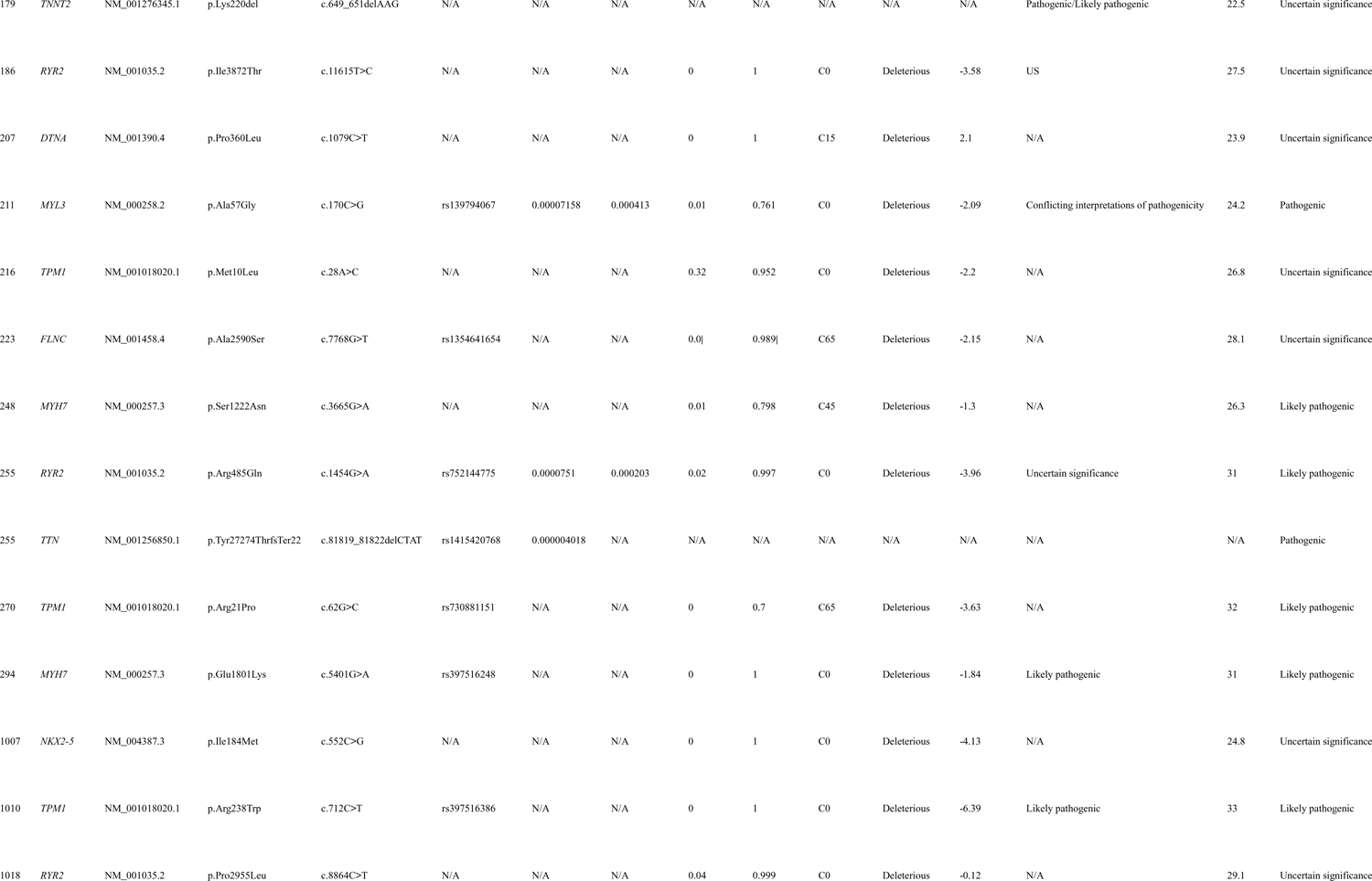
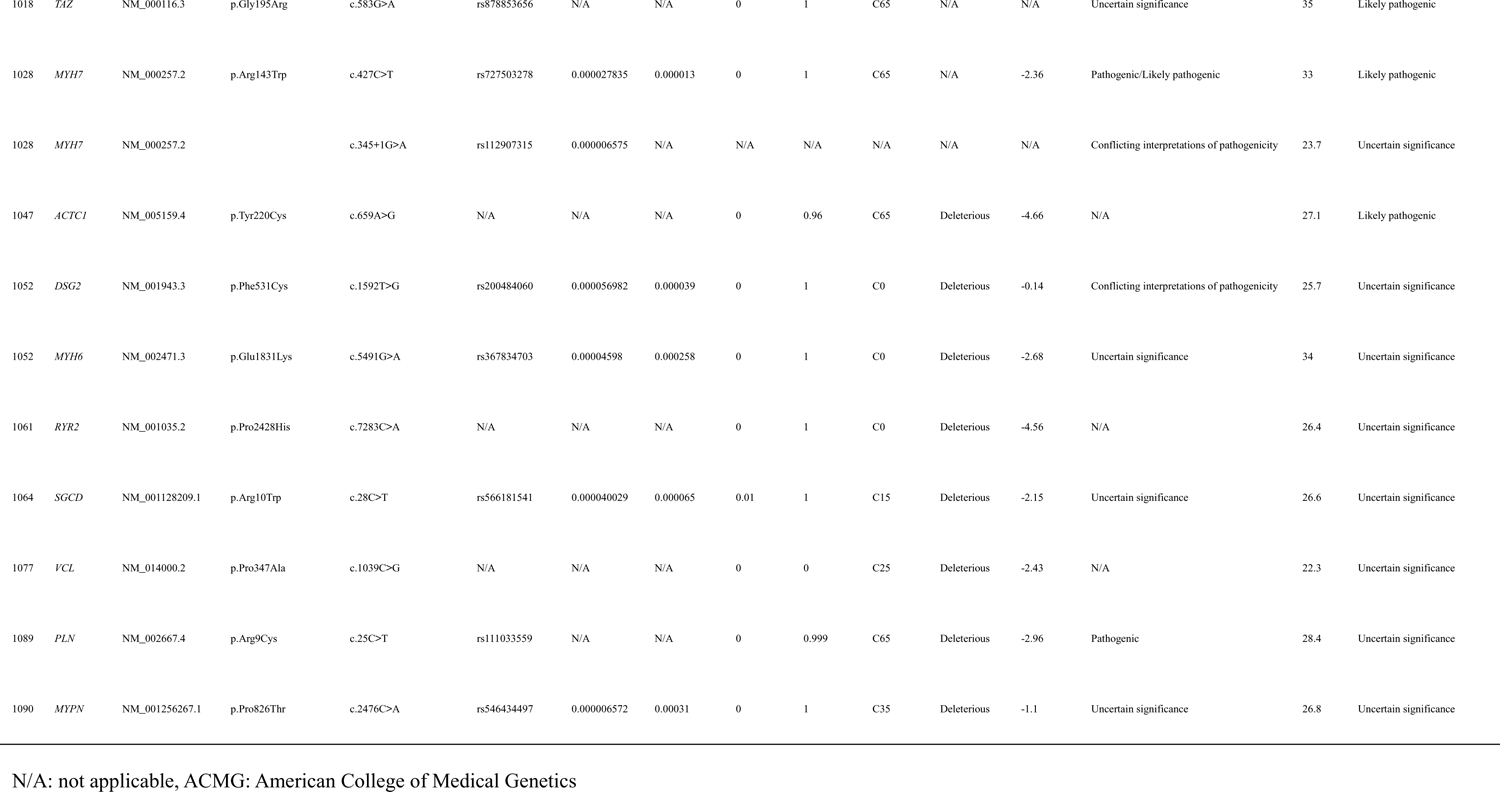
Identified variants in each patient.

To elucidate the genotype–phenotype correlation, patients were divided into six groups according to gene function: 1) sarcomere group, comprising patients with sarcomere gene variants *(ACTC1, MYH6, MYH7, MYPN, TNNC1, TNNI3, TNNT2, TPM1, and TTN);* 2) cytoskeleton group, comprising patients with mitochondrial disease-associated gene variants *(DES, DTNA, FLNC, SGCD, and VCL);* 3) ion channel group, comprising patients with ion channel gene variants (*RYR2*); 4) desmosomal group, comprising patients with desmosomal gene variants *(DSG2 and PKP2);* 5) nuclear envelope group, comprising patients with heart development gene variants *(EMD and LMNA);* and 6) other variants. Variants in genes associated with the sarcomere, cytoskeleton, ion channel, and desmosomal and nuclear envelope groups accounted for 46.0% (n = 23), 12.0% (n = 6), 12.0% (n = 6), 8.0% (n = 4), and 4.0% (n = 2), respectively, for all patients.

Almost all baseline data between individuals with and without sarcomere gene variants did not reveal any differences, except for details regarding diagnostic chance; patients with sarcomere gene variants had fewer symptoms and high fetal screening (Table 3). Most patients with variants in the *MYH7* gene were diagnosed younger than the other patients (Table S3).

**Table 3.**
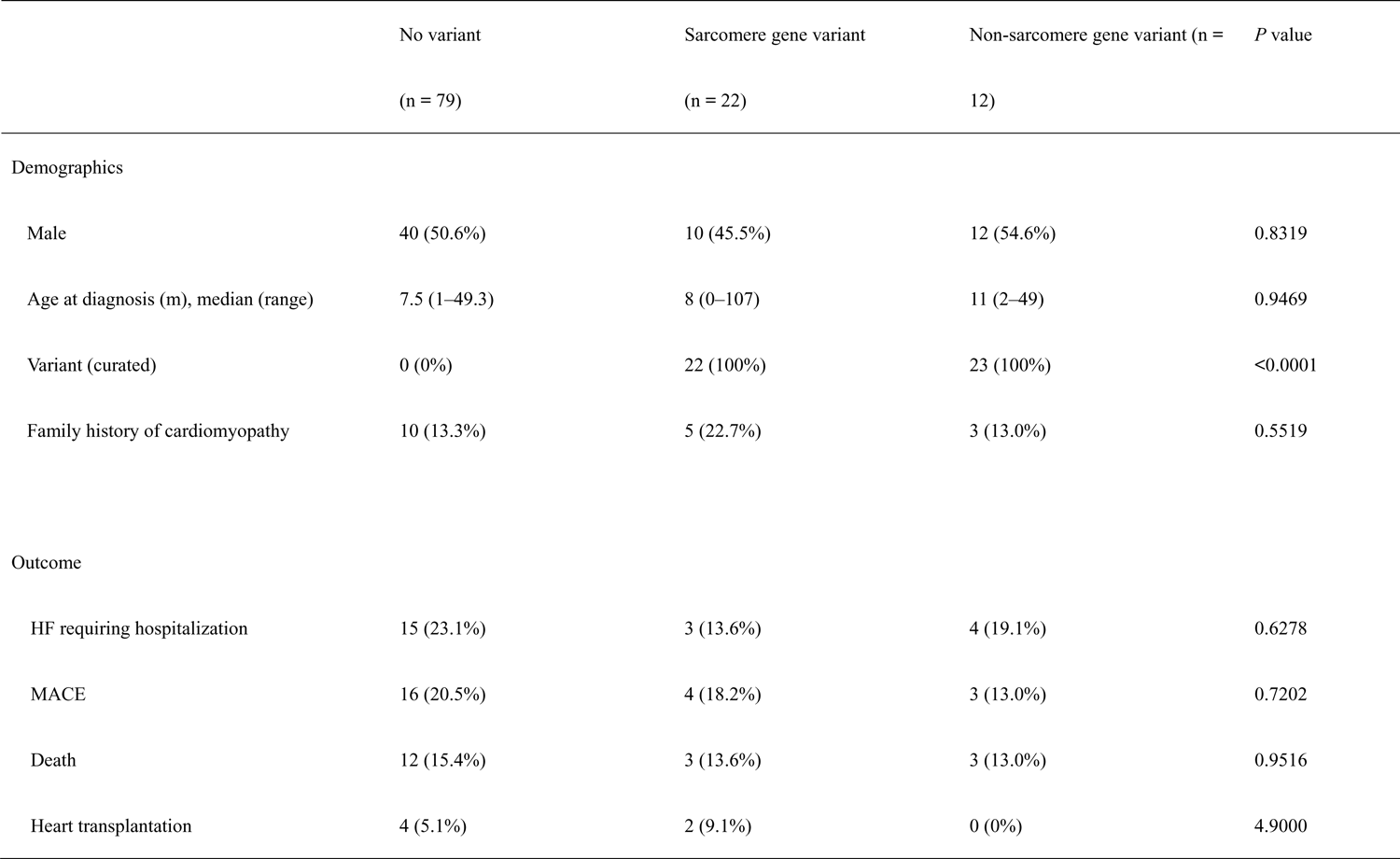
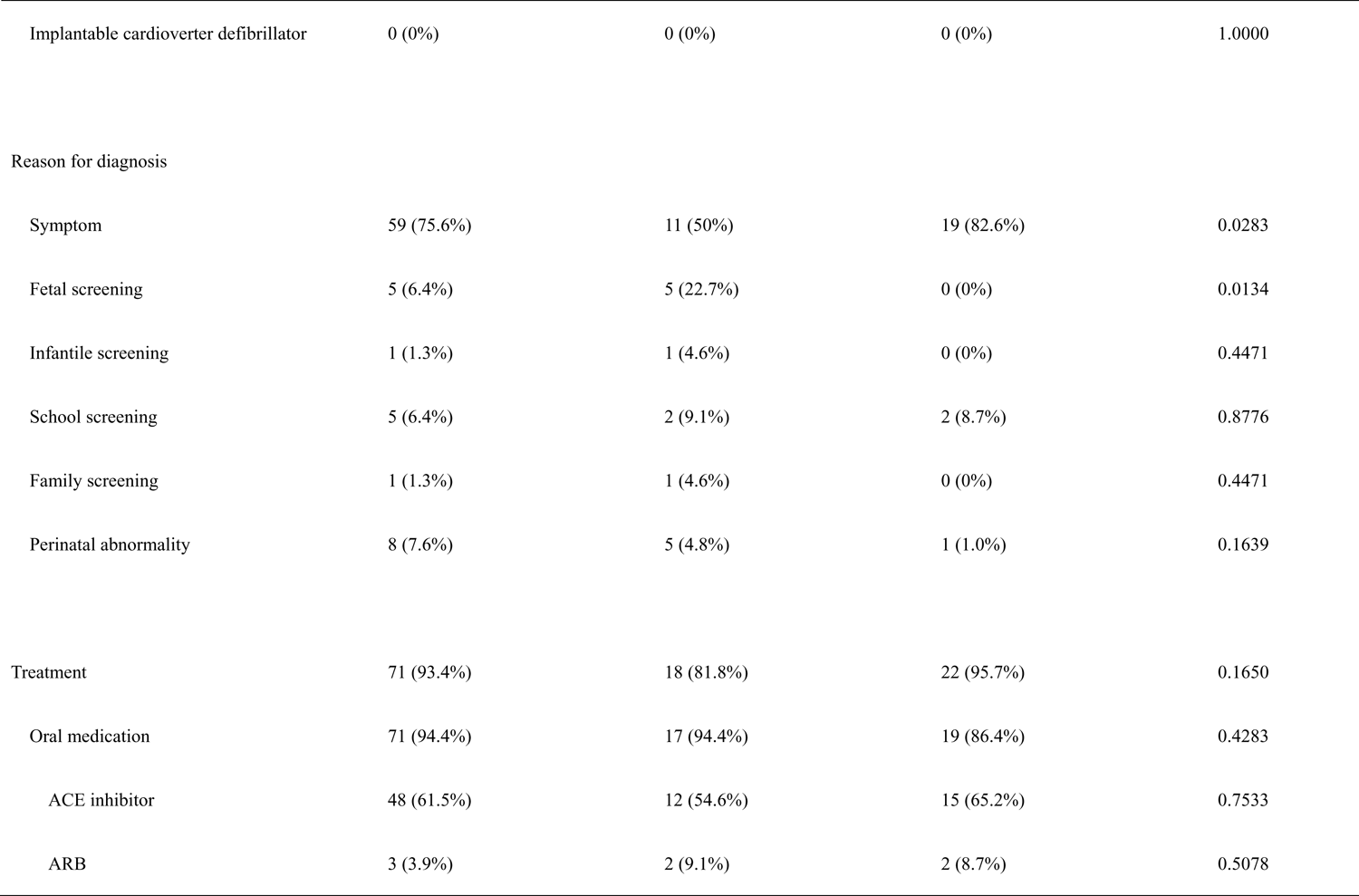
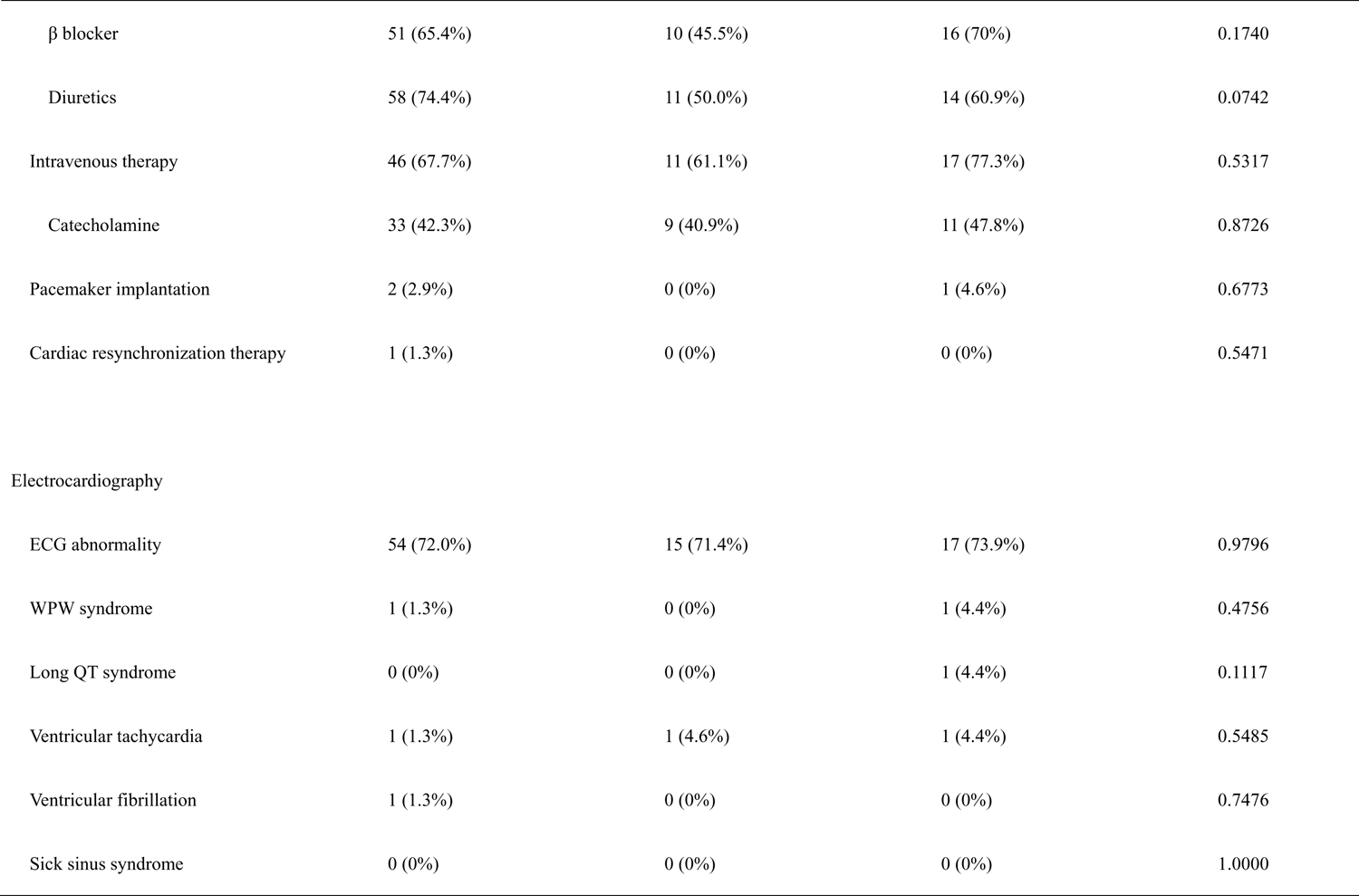
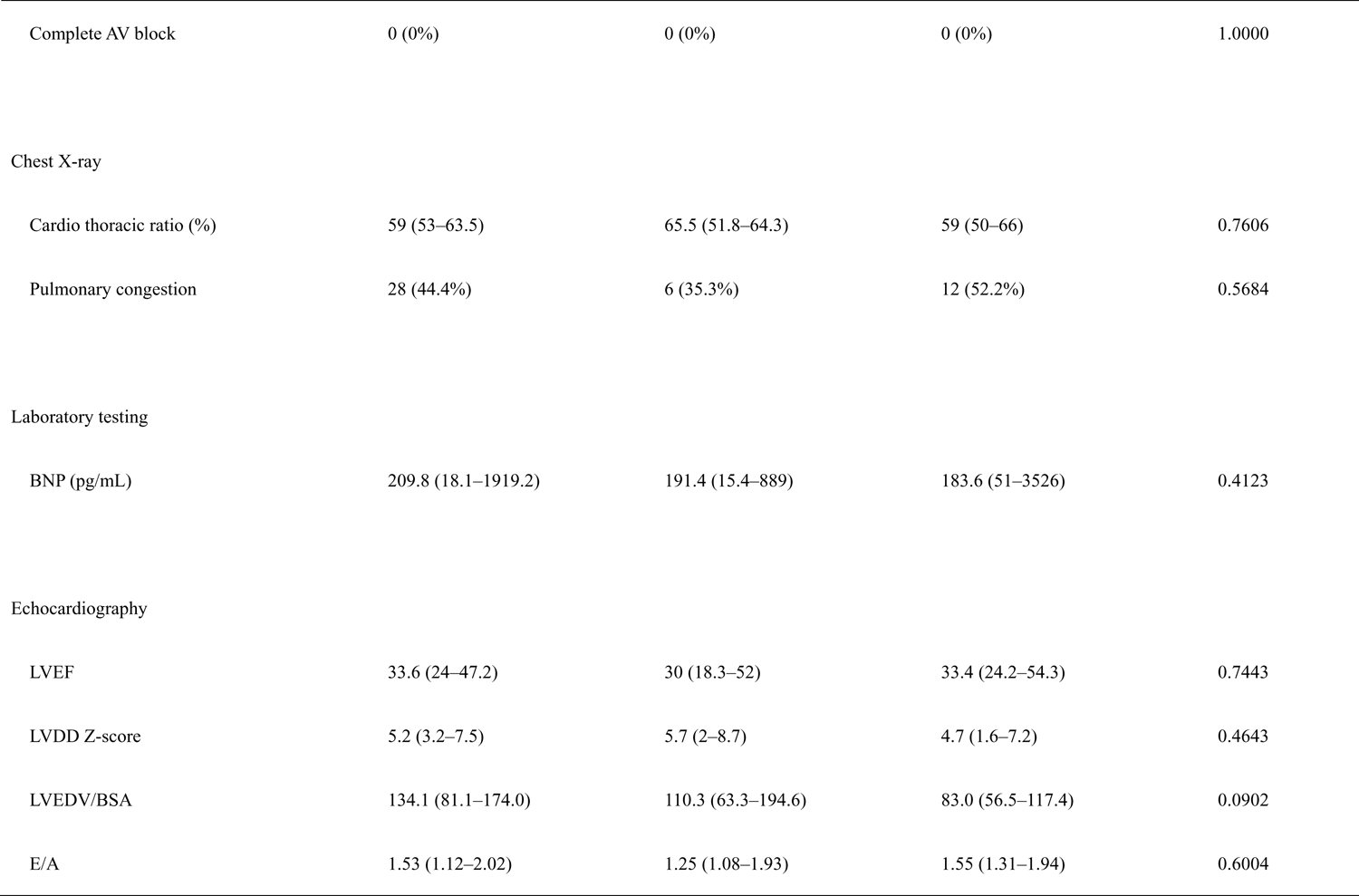
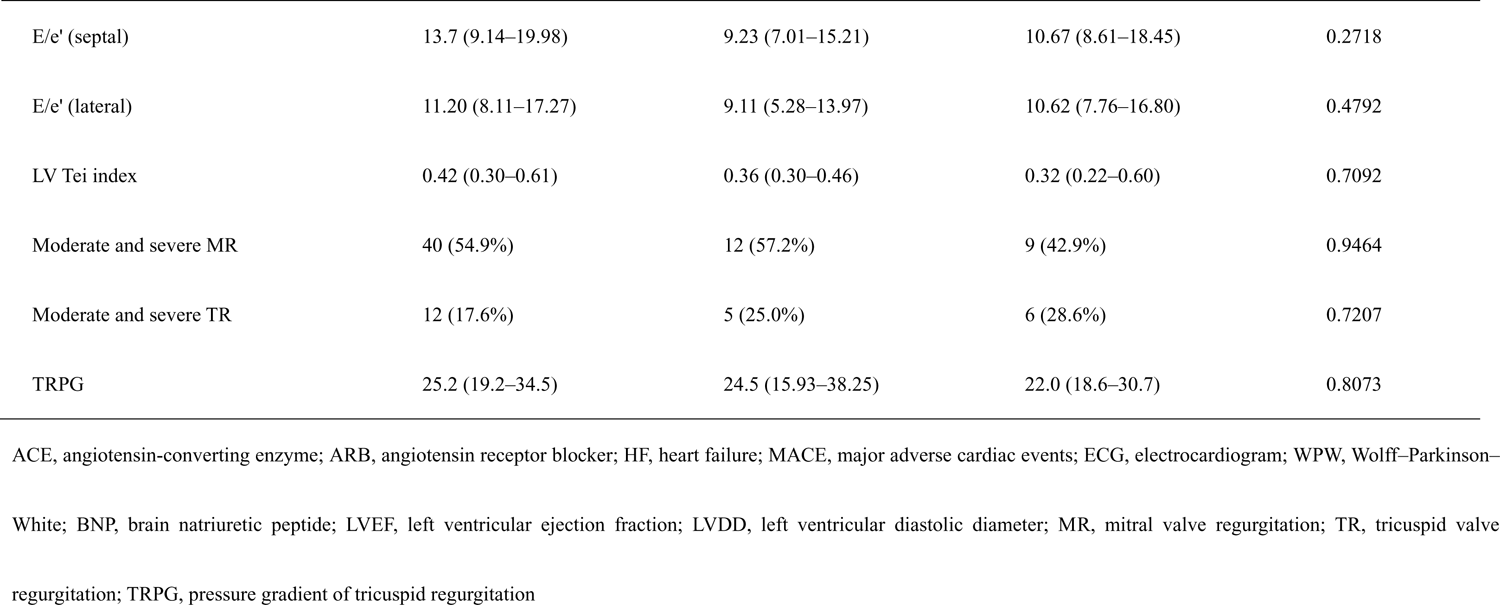
Baseline characteristics stratified by the presence of the sarcomere gene variant.

Of note, a novel variant in the *CASZ1* gene (NM_001079843.2 c.3356G>A, p. Trp1119Ter) that was identified in our cohort (Table 2). A variant in the *MYL3* gene (NM_000258.2 c.170C>G, p. Ala57Gly), which was previously reported in several patients with hypertrophic cardiomyopathy, was identified in a patient with DCM (Table 2).

Interestingly, the sequential change of LVEF and LVDD Z-score between baseline and follow-up was specifically characterized according to gene variants. During the observation period, the LVEF at follow-up in patients with sarcomere gene variants remained unchanged (31.4 [17.2– 53.0] to 39.8 [20.0–62.0], *P* = 0.1913), whereas the LVEF at follow-up in patients with negative gene variants and non-sarcomere gene variants was significantly increased compared with that of baseline (33.4 [24.0–46.2] to 47.8 [28.3–61.7] and 33.6 [27.0–55.6] to 54.1 [35.0–69.5], *P* = 0.003 and *P* = 0.047, respectively) (Figure 2). Similarly, the LVDD Z-score at follow-up in patients with sarcomere gene variants and non-sarcomere gene variants remained unchanged during the observation period (5.35 [1.88–8.40] to 3.11 [−1.24–4.54], *P* = 0.117), whereas the LVDD Z-score at follow-up in patients with negative gene variants was significantly increased (5.37 [3.27–7.74] to 4.27 [2.51–7.63], *P* = 0.006) (Figure 2).

**Figure 2.**
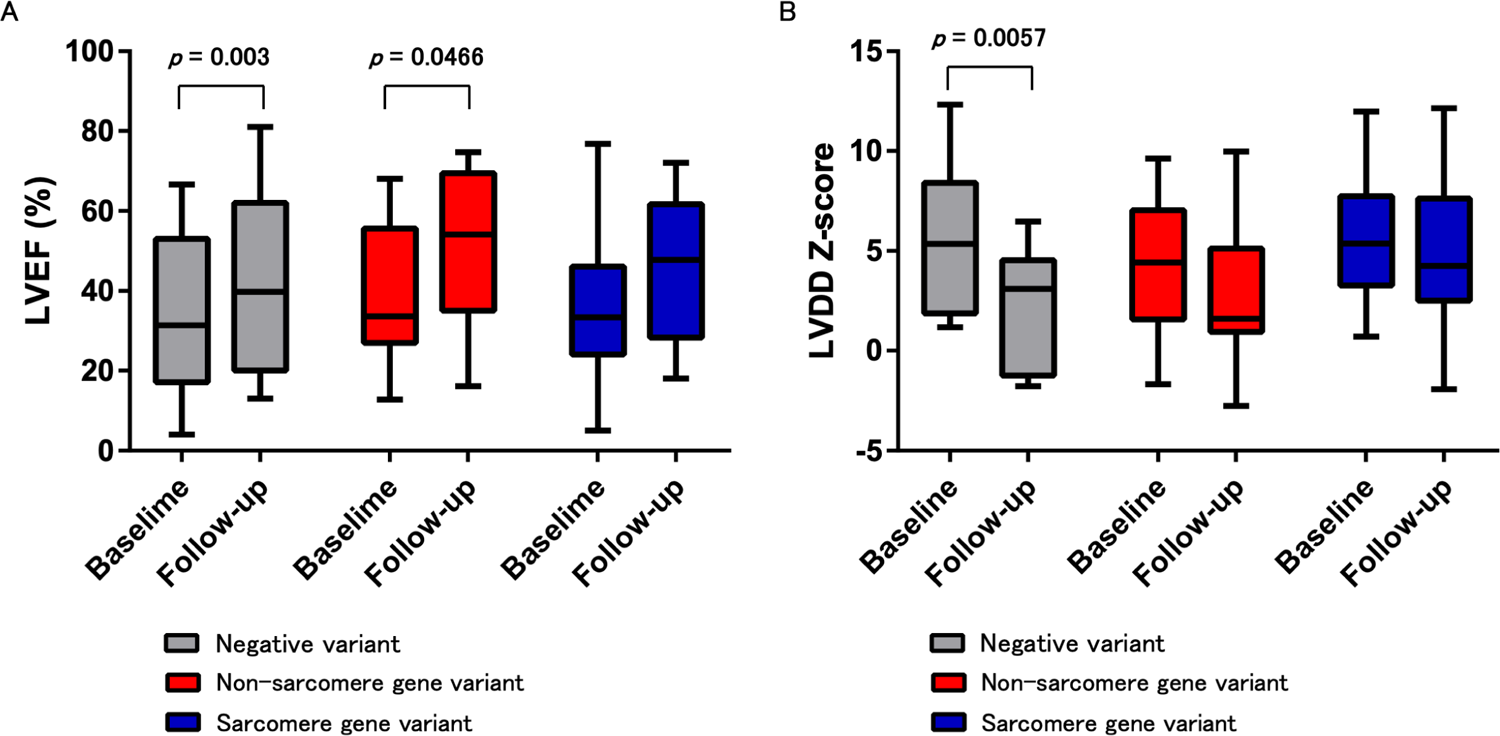
Comparison of serial echocardiographic data among patients according to variants. (A) left ventricular ejection fraction and (B) left ventricular diastolic diameter Z-score.

LVRR at 1 year after the initial echocardiogram at baseline occurred in 47.5% of all patients, 45.0% of the variant-positive group and 50.0% of the variant-negative group; however, the statistical difference was not significant because of the small number of patients (*P* = 0.483) (Figure 3). Differences were also noted in LVRR among the functional gene groups (Figure 3). The worst response was observed in the sarcomere gene group (43%), followed by the non-sarcomere gene group (46%); however, the statistical difference was not significant owing to the small number of patients (*P* = 0.942). No significant differences were found in the LVRR between the pharmacological therapies (data not shown).

**Figure 3.**
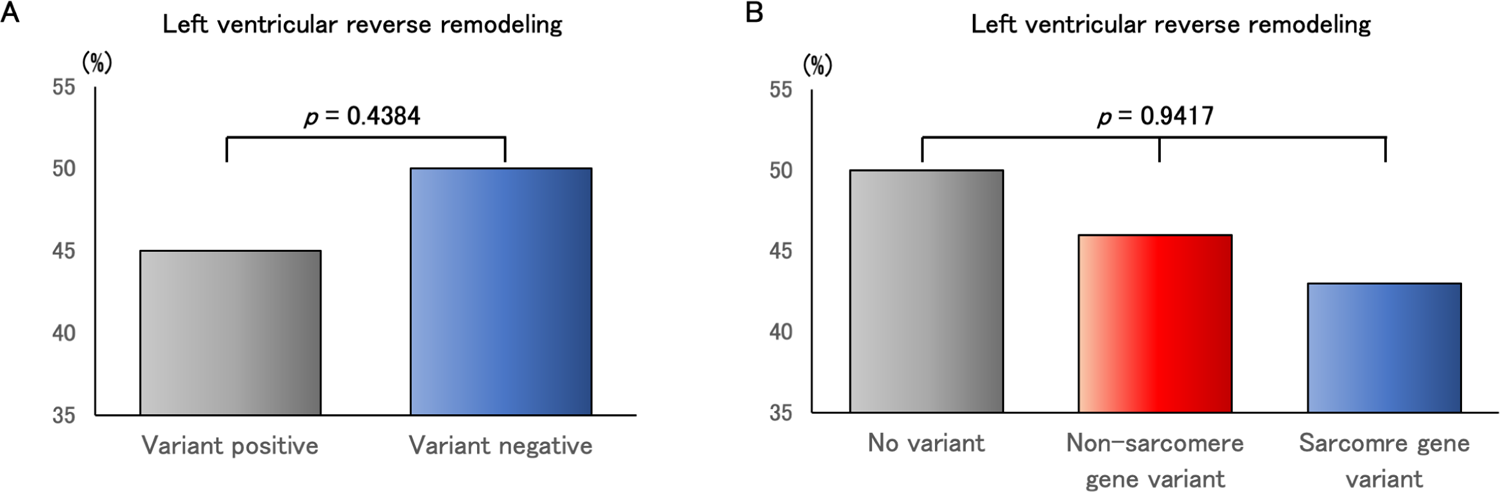
Left ventricular reverse remodeling rate. Left ventricular reverse remodeling rate at the last follow-up in variant-positive versus variant-negative patients with dilated cardiomyopathy (DCM) (A) and the functional gene group in variant-positive patients with DCM (B).

### Clinical outcomes

Among patients with DCM, 22 (17.8%) were hospitalized (Table 1). A total of 111 patients (90.2%) received any treatment. Particularly, oral diuretics, ACE inhibitor, β-blocker, and intravenous catecholamines were administered more frequently.

Adverse events were noted in 23 patients (18.7%), 18 of whom (14.6%) died due to cardiac death, and 6 patients (4.9%) underwent heart transplantation (Table 1). None of the patients had LV assist device implantation or appropriate ICD shock. Figure 4 illustrates the event-free survival from the diagnosis of DCM. Survival rates at 1, 5, and 10 years after diagnosis were 90%, 80%, and 80%, respectively (Figure 4A). Freedom from MACE at 1, 5, and 10 years after diagnosis was 90%, 75%, and 70%, respectively (Figure 4B).

**Figure 4.**
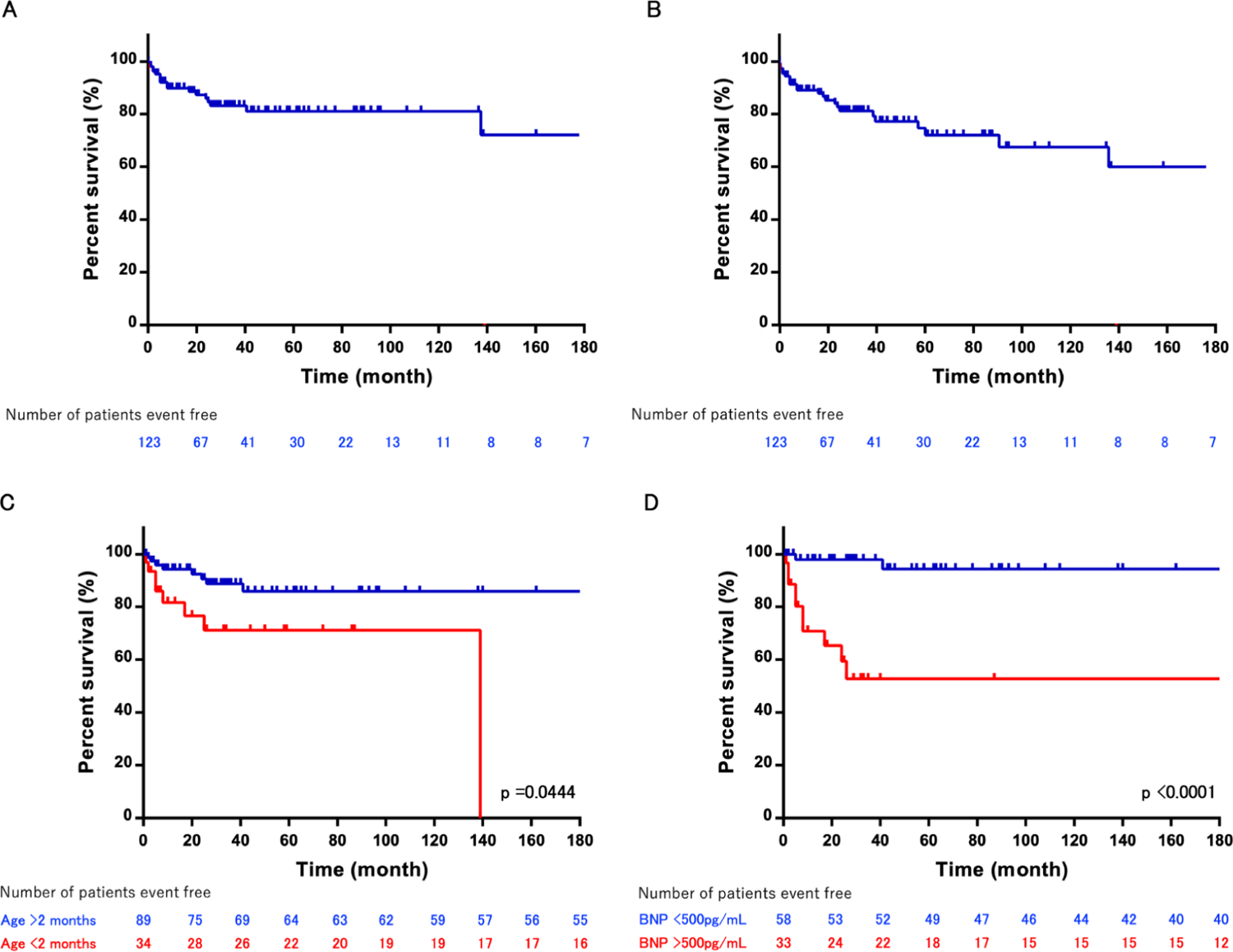
Kaplan–Meier analysis, time from diagnosis to life-threatening cardiac outcome. Event-free survival (A) and freedom from major adverse cardiac events (MACE) (B) in patients with DCM. Event-free survival in patients with DCM (C) according to age (D) and plasma BNP levels. Kaplan–Meier curves were compared using the log-rank test.

Early infantile patients with DCM had a poorer prognosis than older pediatric patients (*P* = 0.044) (Figure 4C). High plasma BNP levels (more than 500 pg/mL) were independent risk factors for survival (odds ratio 7.32; 95% confidence interval 1.56–54.20; *P* = 0.0010) (Table 4 and Figure 4D). No significant differences in mortality or MACE were observed according to gene function (*P* = 0.6653 and *P* = 0.5787, respectively) (Figure S1). The areas under the curve were 0.7646 for BNP to predict survival.

**Table 4.**
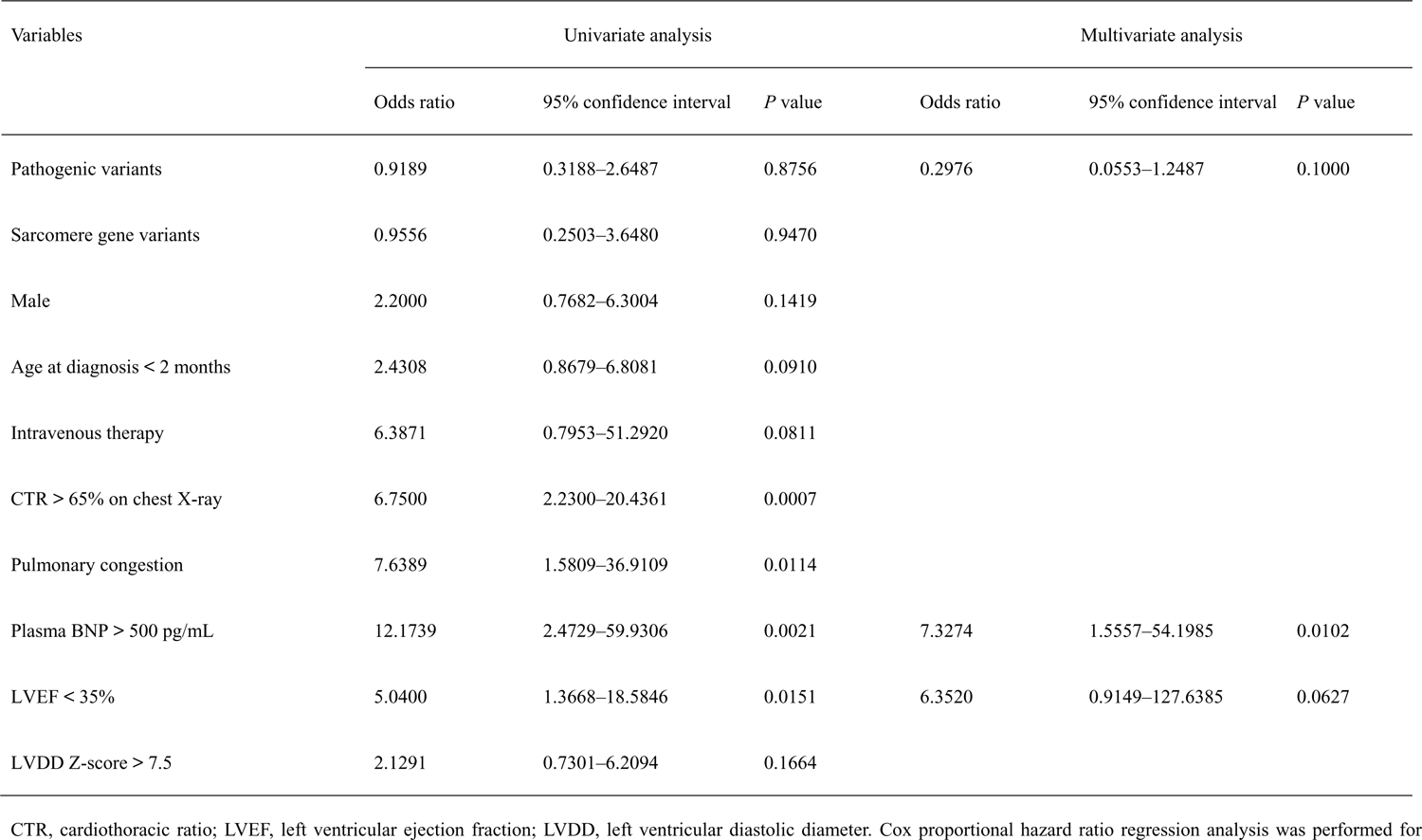
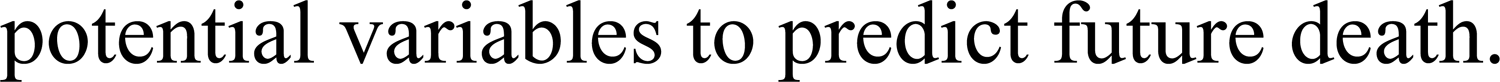
Variables associated with death.

| Variables | Univariate analysis |  |  | Multivariate analysis |  |  |
| --- | --- | --- | --- | --- | --- | --- |
|  | Odds ratio | 95% confidence interval | *P* value | Odds ratio | 95% confidence interval | *P* value |
| Pathogenic variants | 0.9189 | 0.3188–2.6487 | 0.8756 | 0.2976 | 0.0553–1.2487 | 0.1000 |
| Sarcomere gene variants | 0.9556 | 0.2503–3.6480 | 0.9470 |  |  |  |
| Male | 2.2000 | 0.7682–6.3004 | 0.1419 |  |  |  |
| Age at diagnosis < 2 months | 2.4308 | 0.8679–6.8081 | 0.0910 |  |  |  |
| Intravenous therapy | 6.3871 | 0.7953–51.2920 | 0.0811 |  |  |  |
| CTR > 65% on chest X-ray | 6.7500 | 2.2300–20.4361 | 0.0007 |  |  |  |
| Pulmonary congestion | 7.6389 | 1.5809–36.9109 | 0.0114 |  |  |  |
| Plasma BNP > 500 pg/mL | 12.1739 | 2.4729–59.9306 | 0.0021 | 7.3274 | 1.5557–54.1985 | 0.0102 |
| LVEF < 35% | 5.0400 | 1.3668–18.5846 | 0.0151 | 6.3520 | 0.9149–127.6385 | 0.0627 |
| LVDD Z-score > 7.5 | 2.1291 | 0.7301–6.2094 | 0.1664 |  |  |  |

CTR, cardiothoracic ratio; LVEF, left ventricular ejection fraction; LVDD, left ventricular diastolic diameter. Cox proportional hazard ratio regression analysis was performed for
potential variables to predict future death.

## DISCUSSION

Genetic research has made significant strides in elucidating the genetic contributions to cardiomyopathy; however, these studies are largely limited to adults. It is critical to evaluate the genetic structure of pediatric-onset DCM. We established a cohort of 123 pediatric patients with DCM. First, we identified the age at diagnosis of patients and predicted risk factors for survival. Second, 35% of pediatric cases had clinically actionable variants, with the most common genes not being shared between pediatric and adult cases. Third, we evaluated the genetic structure of pediatric DCM and identified LVRR based on the genetic status.

### Genetics

In this study, a retrospective analysis of genetic testing for cardiomyopathy in the largest pediatric cohort with genetically evaluated primary DCM identified disease-causing variants in 35% of patients. In this proband, the detection rate of causal variants was consistent with the results from adult DCM studies and other pediatric cohorts.^21–23^ Additionally, genetic studies that identified variants in pediatric DCM patients revealed age-related differences in the expression of phenotypes based on the affected genes.^24,25^

The high prevalence of variants in sarcomere genes contrasts with adult data, and these variants are more evenly distributed among genes encoding cytoskeletal, nuclear, mitochondrial, and calcium-handling proteins.^26^ There are no hotspots or repetitive variants, and a high prevalence of private variants has been reported. While *LMNA*, *RBM20*, and *PLN* are common in adults, *RBM20* variants were noted in 4% of cases and *PLN* variants in 2% of cases. However, *LMNA* variants were not observed in this study.

Variants in *MYH7* and *RYR2* account for 26% of the disease-causing variants in our cohort and are frequently identified in other pediatric DCM cohorts. Patients with *MYH7* variants all exhibited symptoms in infancy, whereas patients with *RYR2* variants demonstrated symptoms at two different periods. *MYH7* variants have been reported in only 3%–4% of adult-onset DCM; however, in this pediatric cohort, 10% of patients had *MYH7* variants.^27,28^ *MYH7*-related DCM features include an early onset which corroborates with the characteristics observed in our cases.^29^

Among variant-positive pediatric DCM cases, *TTN* truncating variants were observed in two patients (4%), which is a lower proportion than that reported in previous studies.^6,30,31^ Several reports have indicated an association between *TTN* variants and pediatric DCM.^30,31^ *TTN* truncating variants were present in 9% of variant-positive pediatric DCM cases.^6^ Pediatric cases developing DCM in their teens had a fourfold higher likelihood of possessing *TTN* variants compared with those discovered at a younger age. Our data of a lower prevalence of *TTN* truncating variants may reflect an age difference in our study compared to another previous research.

Furthermore, two novel genes associated with the disease were identified: a variant in the *CASZ1* gene (NM_001079843.2 c.3356G>A, p. Trp1119Ter) and *MYL3* gene (NM_000258.2 c.170C>G, p. Ala57Gly). *CASZ1* is a para-zinc-finger transcription factor that is required for vertebrate heart development.^32^ Only 3 variants have been described (Leu38Pro, Lys351Term, and Val815Profs*15).^33–36^ All of these are loss-of-function variants located in the coding region and may be associated with DCM. *CASZ1* variants may result in haploinsufficiency due to nonsense-mediated decay, which plays an important role in the pathological mechanism of DCM.^33^ A variant in the *MYL3* gene is a sarcomere gene that was previously reported in several patients with hypertrophic cardiomyopathy and was identified in a patient with DCM.

### LVRR

In this study, we investigated the genetic basis of Japanese patients with DCM and explored the association between genotypes and phenotypes and revealed an association between LVRR and the genotype. LVRR is known to occur in approximately 40% of patients with DCM under medical treatment.^11,37^ LVRR was recently observed with optimal oral treatment in 5 cases of Japanese pediatric early onset DCM.^38^ While achieving LVRR is associated with a favorable prognosis in patients with DCM, specific genotypes involved in LVRR are unknown in pediatric patients.^11^

We observed that LVRR occurred more frequently in variant-negative patients than in variant-positive patients, and a strong inverse correlation was noted between variants in sarcomere genes and LVRR. Recent studies supported this finding that, compared with the favorable prognosis in the variant-negative group, the occurrence of LVRR is lower in the sarcomere and desmosome gene groups.^10,39^

Other groups have studied the incidence of LVRR in patients with DCM using different definitions. These criteria were based on LVEF and LVDD improvement.^11,40^ Our study underscored on pediatric patients, and LVDD varies with age. Thus, LVDD was deemed inappropriate, and this criterion was not used. Although significant differences were observed in the baseline and follow-up values of LVEF and LVDD based on the genetic type, no significant difference was found in LVRR because of the limited sample size. Future studies should accumulate more cases for further validation.

### Outcome

As a result, the distribution of onset age dominated in the younger age group, especially under 1 year old. In Japan, heart disease screening is conducted in schools to determine those who need further examination and to manage school activities for those with underlying heart diseases.^41^ Some studies have reported the utility of school-based screening programs, which are beneficial for early diagnosis of arrhythmia and cardiomyopathies. In patients with LVNC, school screening of patients accounted for 41.9% of diagnosed patients in the overall pediatric patients and 59.5% of school children.^42^ However, in this study, while there appears to be a minimal peak in cases of DCM that are likely detected in the school-age period, it is not as pronounced as observed with LVNC. This is believed to be due to the differences in the proportion of cases with clinical symptoms, as DCM patients tend to exhibit clinical symptoms at a younger age, whereas patients with LVNC are asymptomatic and incidentally diagnosed during school screenings.

In this study, the survival rates at 1 and 5 years were 90% and 80%, respectively. Freedom from death or heart transplantation rates at 1 and 5 years after diagnosis were 90% and 75%, respectively. A recent Japanese cohort study reported that the survival rates at 1 and 5 years after diagnosis were 81% and 75%, respectively, and freedom from death or heart transplantation rates at 1 and 5 years after diagnosis were 76% and 64%, respectively.^43^ The difference in outcomes between both studies can be attributed to the fact that our study targeted patients from 2014 to 2023, while the recent Japanese cohort study included patients from 1990 to 2014, reflecting differences in medical practices over different time periods. These tendencies of the survival rates or heart transplantation were similar to the data from the U.S. study (84% and 76% at 1 and 5 years, respectively).^17^ In the Australian cohort study, the survival rates or heart transplantation at 1 and 5 years after diagnosis were 74% and 65%, respectively.^14,44^ The Australian study included patients with clear evidence of myocarditis, whereas they were excluded from the present study. The long-term incidence of death or heart transplantation in the pediatric population is markedly higher than that in the adult population, as reported in the U.S., Australia, and Japan.^4,45–47^ This may be attributed to genetic causes; for instance, *TTN* variants were predominantly found in adult patients but were rare in pediatric patients in our study. Our report shows a better prognosis than previously reported results, which emphasizes the importance of LVRR.

### Risk factors

In our multivariate analysis, elevated plasma BNP levels were identified as an independent risk factor for survival. Additionally, a younger age may be a risk factor for survival. In pediatric cohorts, similar to N-terminal-pro-BNP, reduced LV function, older age, familial cardiomyopathy, and an increase in heart rate have been recognized as risk factors for survival in pediatric DCM patients.^14,17,43,44,48,49^ In a multivariate analysis, Meulen et al. reported that N-terminal-pro-BNP was the sole independent predictor for adverse outcomes.^50^ Mori et al. found that a reduced LV function at baseline was associated with an increased risk of death or heart transplantation in Japan, including in Australia and the U.S., supporting the results of our univariate analysis.^14,17,44^ Previous reports indicated that older age and familial cardiomyopathy were linked to an increased risk of death or heart transplantation.^14,17,44,48^ However, in our study, older age and familial cardiomyopathy were not significant risk factors. The reasons for this difference remain unclear but may be multifactorial, including factors such as age distribution, racial differences, and differences in the study period. In another study using univariate analysis, an elevated heart rate was reported to be associated with an increased risk of death and the combined outcome of death or heart transplantation.^49^ Unfortunately, we did not thoroughly evaluate heart rate at baseline and follow-up in patients with DCM. Further research is required to validate these results.

### Study limitations

There were limitations in determining whether segregation analysis or de novo occurrence could explain certain variants because some parental samples were not available. Some cases included in this study were registered over 5 years ago, and new treatments have been developed, potentially altering the outcomes. The selection of an NGS panel targeting genes known to be associated with the cardiac phenotype and development may have resulted in the oversight of unknown variants because information from the entire genome was not obtained. Inadequate follow-up investigations are possible because of missing data in some cases. Given the small sample size, future large-scale studies are necessary for further elucidation of genotype–phenotype correlations. Additionally, phenotype analysis through comprehensive diagnostics involving magnetic resonance imaging and pathological tissue examination would be desirable.

## Conclusions

Our results confirm that pediatric DCM exhibits marked genetic heterogeneity, with a different landscape from adult DCM. Younger patients had predominance in DCM and risk factors for survival. LVRR was not uniform across functional gene groups, opening the door to the adoption of an individualized prediction approach in DCM based on genetic features.

## Data Availability

The data that supports the findings of this study are available in the supplementary material of this article.

## Acknowledgments

The authors wish to acknowledge Hitoshi Moriuchi, Haruna Hirai, and Eriko Masuda for their expert technical assistance.

## Abbreviations

DCM: dilated cardiomyopathy

ECG: electrocardiogram

HF: heart failure

LV: left ventricle

LVRR: left ventricular reverse remodeling

MACE: major adverse cardiac events

NGS: next-generation sequencing

PCR: polymerase chain reaction

BNP: brain natriuretic peptide

